# Sudden death in patients with sleep apnea: a systematic review and meta-analysis

**DOI:** 10.1101/2020.05.06.20093641

**Authors:** Emily S. Heilbrunn, Paddy Ssentongo, Vernon M. Chinchilli, Anna E. Ssentongo

## Abstract

**Background:** Over 1 billion individuals across the globe experience some form of sleep apnea, and this number is steadily rising. Obstructive sleep apnea (OSA) can negatively influence one’s quality of life and potentially increase the risk of mortality. However, this association between OSA and mortality has not been comprehensively and thoroughly explored. This meta-analysis was conducted to conclusively estimate the risk of death for all-cause mortality and cardiovascular mortality in OSA patients.

**Study Design:** 4,613 articles from databases including PUBMED, OVID & Joana Briggs, and SCOPUS were comprehensively assessed by two reviewers (AES & ESH) for inclusion criteria. 28 total articles were included, with 22 of them being used for quantitative analysis. Pooled effects of all-cause mortality, cardiac mortality, and sudden death were calculated by utilizing the metaprop function in R Statistical Software and the random-effects model with appropriate 95% confidence intervals.

**Results:** Analysis on 42,032 individuals revealed that those with OSA were twice as likely to die from cardiac mortality compared to those without sleep apnea (HR= 1.94, 95%CI 1.39-2.70). Likewise, individuals with OSA were 1.7 times as likely to die from all-cause sudden death compared to individuals without sleep apnea (HR= 1.74, 95%CI 1.40-2.10). There was a significant dose response relationship between severity of sleep apnea and incidence risk of death, where those with severe sleep apnea were

**Conclusions:** Individuals with obstructive sleep apnea are at an increased risk for all-cause mortality and cardiac mortality. Further research related to appropriate interventions and treatments are necessary in order to reduce this risk and optimize survival in this population.

**Key Messages:** *What is the key question?:* Are individuals with sleep apnea at an increased risk for cardiovascular mortality and sudden death?

*What is the bottom Line?:* Sleep apnea is associated with an increased risk of cardiovascular mortality and sudden death, with a dose response relationship, where those with severe sleep apnea are at the highest risk of mortality.

*Why read on?:* This is the first systematic review and meta-analyses to synthesize and quantify the risk of mortality in those with sleep apnea, highlighting important directions for future research.

*Prospero Registration ID:* **CRD42020164941**

## Introduction

Obstructive sleep apnea (OSA) is a chronic sleep disorder, in which the patient experiences complete or partial obstruction of the upper airway structures [1-2]. This then results in a reduction or complete blockage of airflow during sleep, intermittent hypoxia, and sleep disturbances [3]. Common symptoms of OSA, such as excessive daytime sleepiness, fatigue, heavy snoring, and non-refreshing sleep have the potential to play an influential role in one’s quality of life (QOL) [4]. OSA also increases one’s risk of developing several cardiovascular-related comorbidities, including heart failure, arrhythmias, and coronary artery disease [5]. Those with apnea-hypopnea index (AHI) > 36 have a higher risk of all-cause mortality in comparison to other AHI scores (HR = 3.30; 95% CI 1.74-6.26) [6]. Patients with obstructive sleep apnea who are classified as moderate (AHI 15-30) to severe (AHI >30) have an increased risk of many adverse outcomes, specifically all-cause mortality [7]. Therefore, we hypothesize that obstructive sleep apnea has the potential to act as a predictor for cardiac events and general mortality. As a result of this, the risk of mortality in patients diagnosed with obstructive sleep apnea has become a growing and pervasive concern.

OSA is a growing public health concern, with previous studies in the literature reporting an increasing global prevalence that is upwards to 1 billion individuals [8]. Estimates within the United States suggest that approximately 15% of adults are clinically diagnosed with obstructive sleep apnea [9]. There is a large portion of the population who suffer from OSA-like symptoms, without receiving any diagnosis or treatment. This group continues to further the burden placed on the public health and medical systems. Identifying individuals diagnosed with OSA who are considered at-risk for adverse health outcomes can reduce the economic cost and burden on the healthcare system [10].

Existing evidence on the association between OSA and the risk of sudden death is insufficient and inconclusive. Our objective is to explore the association of OSA and the mortality risk. We hypothesize that in patients with OSA, there is an increased risk of all-cause mortality, cardiovascular mortality, and sudden death. To our knowledge, this is the first comprehensive systematic review and meta-analysis that explores the association between OSA and all-cause mortality, cardiovascular mortality, and sudden death.

## Methods

### Patient and Public Involvement

Patients or the public were not involved in the preparation or dissemination of this manuscript.

### Search Strategy and Selection Criteria

We comprehensively searched five databases for articles, including PubMed (MEDLINE), Cochrane, OVID (Healthstar), OVID (Medline), Scopus, and Joanna Briggs Institute EBF Database. We followed the Meta-analysis of Observational Studies in Epidemiology (MOOSE) standards for this research [11]. We identified observational studies published prior to January 1st, 2020 with reported rates of sudden death in individuals with OSA. We searched the citation list of included papers by using the snowballing method, i.e., we screened reference lists of these articles for potential eligibility as well. We did not impose any limitations related to the date of publication, language, study design, or geographical location. We predetermined defined search terms by using Medical Subject Headings (MeSH) and included combinations of “Sleep” AND “obstructive” AND “apnea” AND “sudden” AND “death” OR “sleep” AND “obstructive” AND “apnea” AND “death” OR “apnea” AND “death” AND “sleep.” We initially inputted potential studies into EndNote, which removed all duplicate studies. Two reviewers (ESH and AES) independently screened titles and abstracts of the studies for inclusion eligibility. The inclusion criteria involved studies that report the rates of sudden death in both patients with and without OSA, including information on the incidence and prevalence of the outcome. Excluded studies were those that were not conducted on humans and did not report the prevalence or incidence of the outcome of interest. We excluded review papers, meta-analyses, literature reviews, commentaries, and meeting abstracts as well. We documented all excluded studies with reasons for their exclusion.

### Quality Assessment and Data Extraction

Since all of our studies were nonrandomized observational studies, we used the Newcastle-Ottawa Scale (NOS) for quality assessment [12]. After screening initial articles based on their titles and abstracts, ESH and AES manually screened full-text articles for continued inclusion criteria. In the event of a potential disagreement, a third researcher (PS) was recruited in order to reach a consensus. Appropriate and eligible data were extracted when screening eligible studies, including the full title, year of publication, country of publication, number of participants with and without OSA, median age of participants in both study groups, median BMI of participants with and without OSA, proportion of the sample that was male, proportion of the study sample who smoked, proportion of the study sample with hypertension and diabetes, the number of participants with OSA who died, the number of participants without OSA who died, the number of participants who did not die from both groups, the RR/OR/HR of sudden death, and other findings of interest. Studies not published in English were translated.

### Data Analysis

The primary outcome of interest for this meta-analysis and systematic review was the incidence of all-cause mortality in patients with OSA. There were two secondary outcomes of interest, namely, cardiac mortality and sudden death. We used the metaprop function in R Statistical Software to calculate the pooled effects of all-cause mortality, cardiac mortality, and sudden death. We invoked the random-effects model with a logit transformation of proportions. We applied this random-effects model, regardless of the presence of heterogeneity across studies. We assessed inter-study heterogeneity using the *I*^2^ statistic, which was defined as low (25%), moderate (50%), and high (75%) (*a* < 0.05). We conducted a meta-regression using study-level descriptive statistics of age, sex, race, BMI, smoking status, diabetes, and hypertension as repressors in order to improve the precision of the meta-analysis. We calculated confidence intervals using the Copper-Pearson exact binomial interval method. We investigated potential publication bias via funnel plots and Egger’s test.

## Results

As described in **Figure 1**, we identified a total of 4,613 studies from Scopus, OVID, PubMed, and Joana Briggs International EBF database. We excluded 1,443 studies because they were duplicates, leaving 3,170 studies to explore for inclusion. Based on the title and abstracts, we excluded another 2,893 studies based on titles and abstracts, and another 249 based on full text. This left us with 28 studies for the qualitative analysis and 22 studies for the quantitative analysis [13-40]. Of the studies included in the quantitative analysis, 12 studies were from North America, 5 from Europe, 3 from Asia, and 1 from South America.

**Figure 1:**
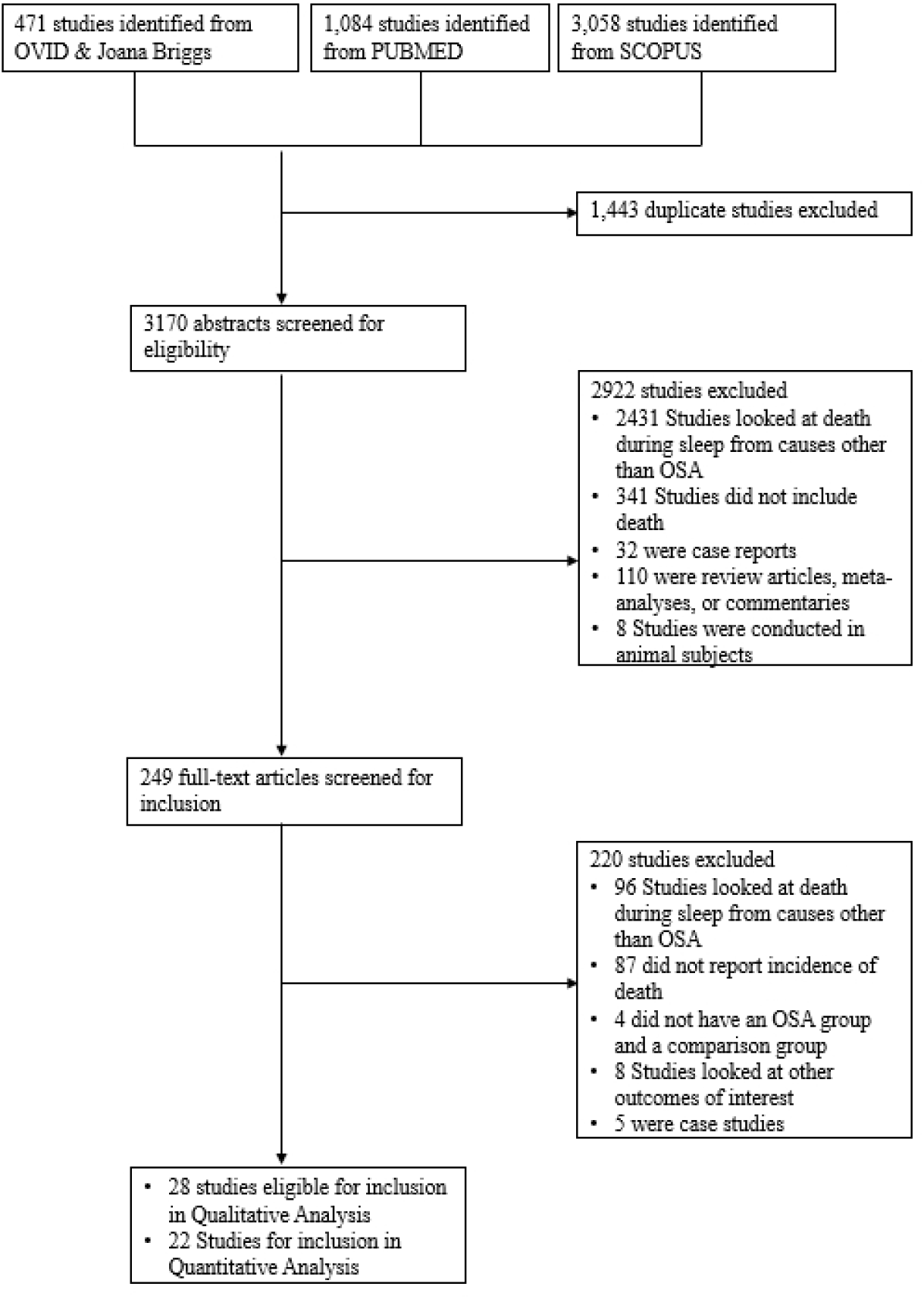
Flow Diagram displaying the studies of inclusion. Of 4,613 studies explored, 28 were deemed eligible for inclusion in the qualitative analysis, and 22 studies reporting on 42,032 individuals were included in the quantitative meta-analyses.

### The Association of OSA and Sudden Death

The hazard ratio of mortality from OSA ranged from 0.80 to 9.20 (**Figure 2**). Those with OSA displayed nearly twice the hazard for sudden death compared to those without OSA (HR: 1.74 95%CI 1.4-2.10). Those in Australia had the highest rate of mortality from OSA (HR= 6.24 95%CI 1.44 -26.98), followed by South America, Europe, Asia, and North America **Figure 2)**.

**Figure 2:**
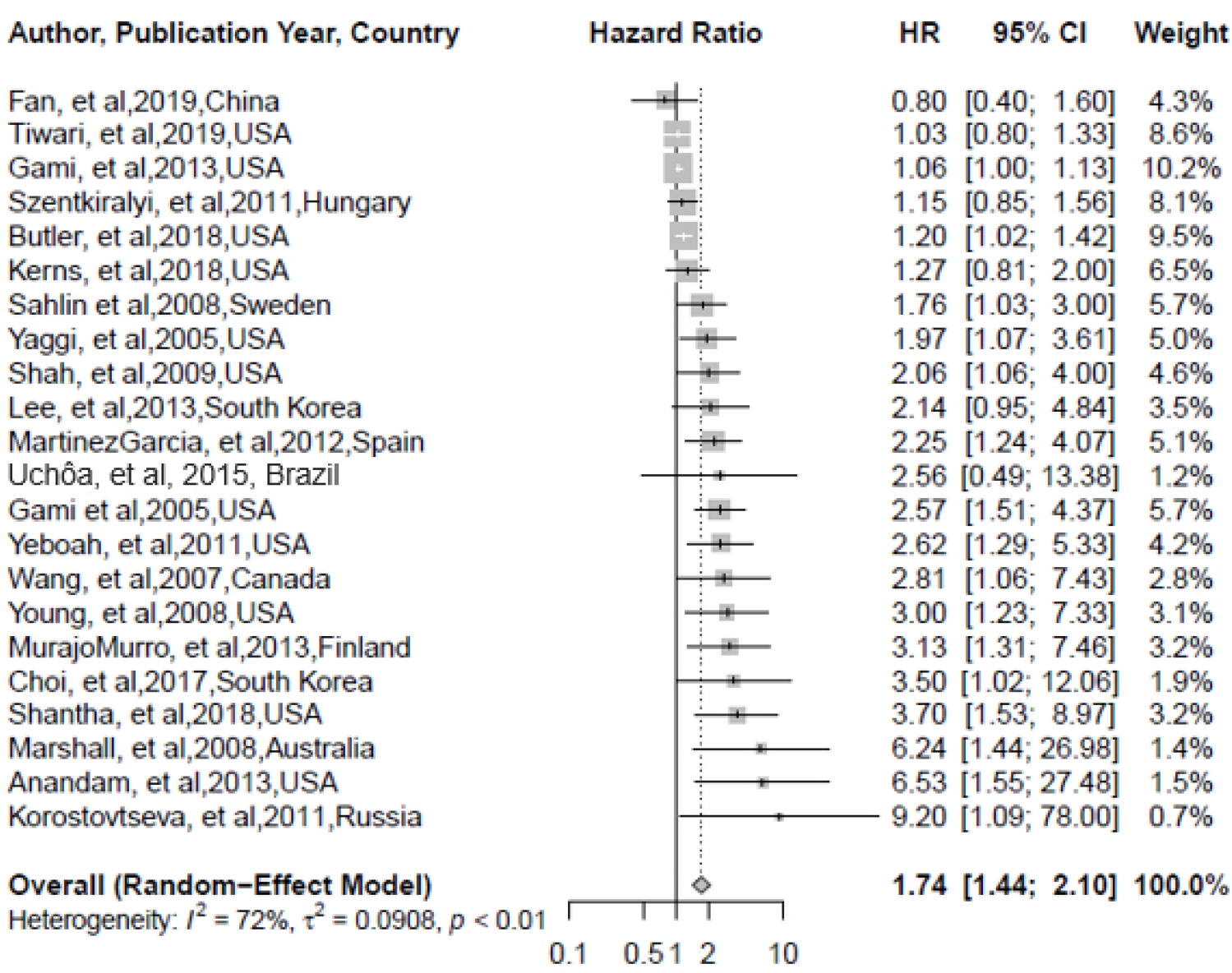
All cause mortality. Individuals with sleep apnea were 1.7 times as likely to die from all-cause mortality compared to those without sleep apnea (HR=1.74, 95%CI 1.44-2.10).

**Figure 3:**
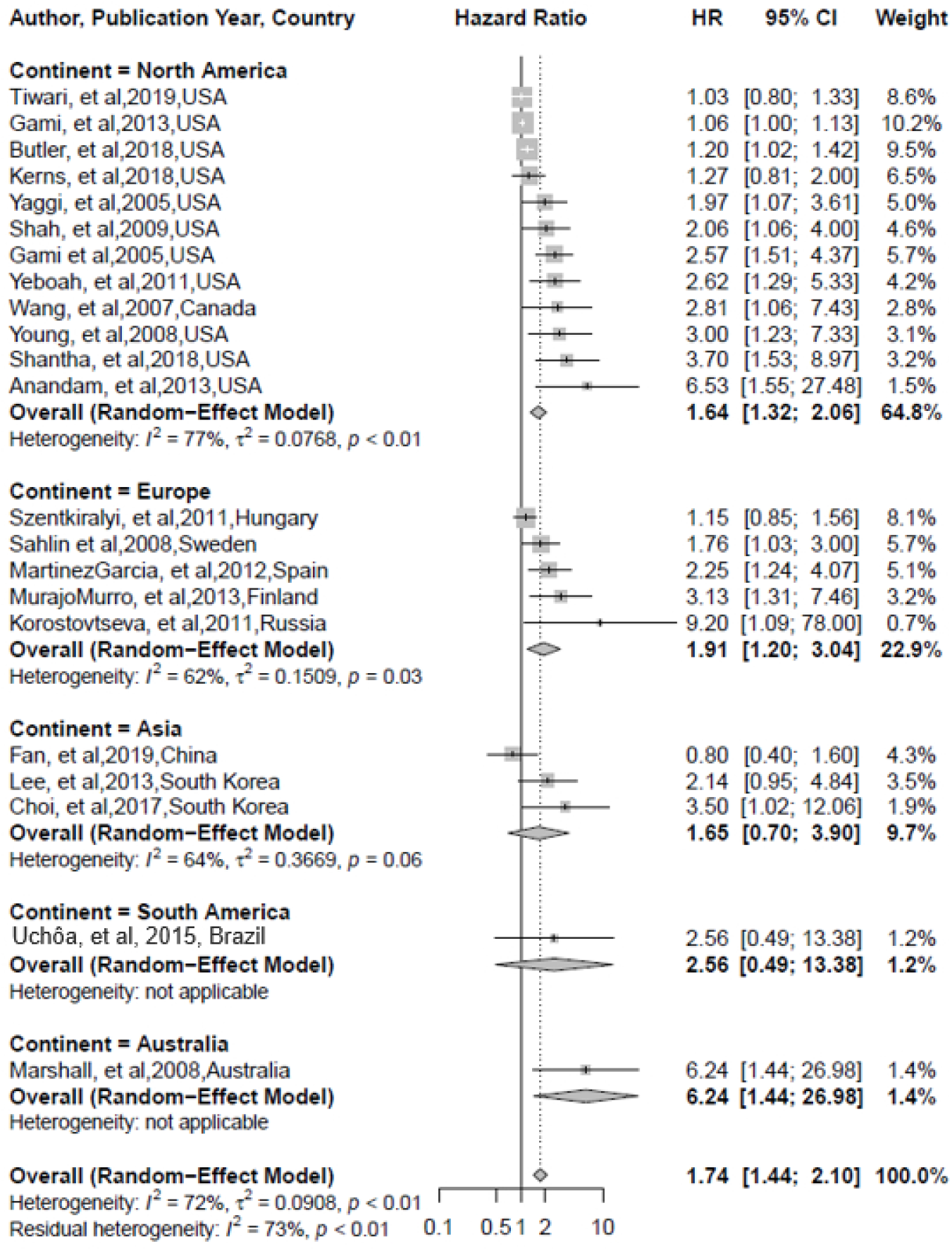
All cause mortality by continent. Although not statistically significant, individuals in Europe with sleep apnea had a higher incidence of mortality compared to those in North America. Australia and South America had the highest incidence of all-cause mortality from sleep apnea, although these continents urgently need more studies.

**Figure 4:**
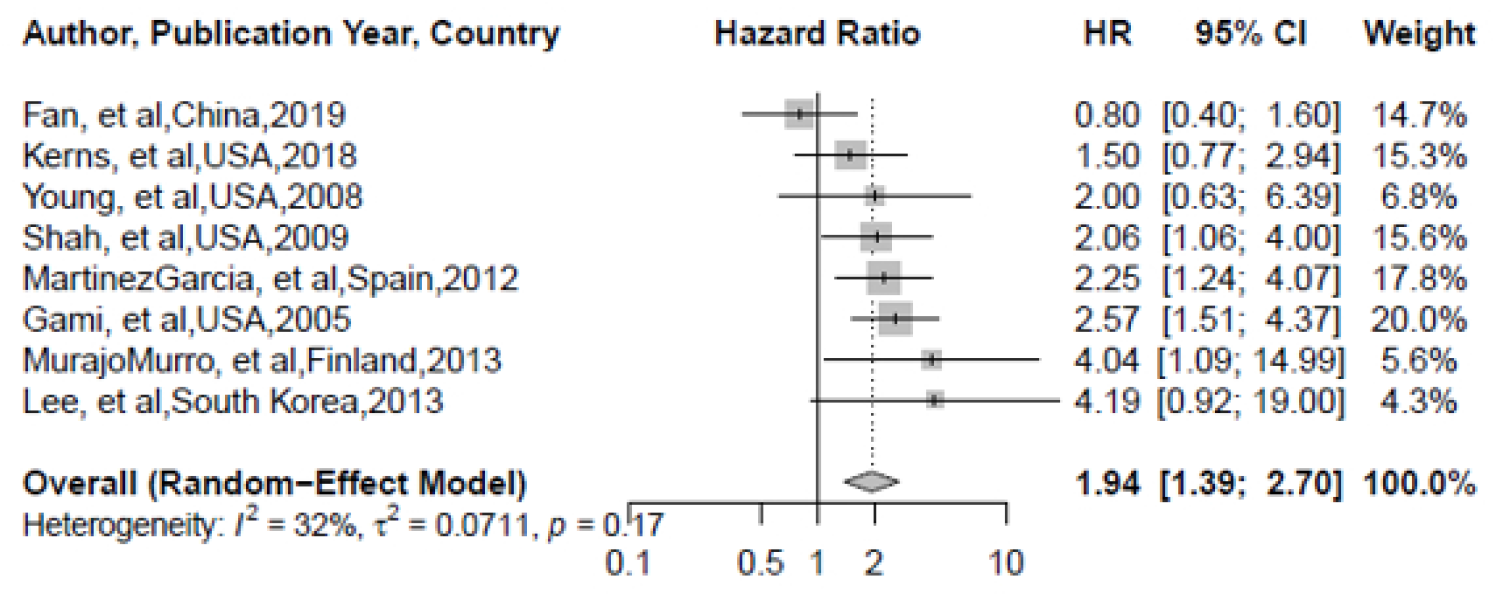
Cardiovascular Mortality. Individuals with sleep apnea were twice as likely to die from a cardiovascular related cause compared to those without sleep apnea (HR: 1.94, 95%CI 1.39-2.70).

### The Association of OSA and Cardiac Mortality

The rate of cardiovascular mortality ranged from 0.80 to 4.19 (**Figure 2)**. Overall, those with OSA displayed nearly twice the hazard for cardiovascular mortality compared to those without OSA (OR= 1.94, 95%CI 1.39-2.70). Only two studies reported the HR of sudden death. The HR for sudden death ranged from 1.06 to 3.28.

### Dose-response Effect of OSA of Sudden death

Five studies reported the HR of all-cause mortality depending on the severity of OSA. There is a dose response relationship between the relationship of OSA and sudden death. The hazard ratio for mild was 1.16 95% CI 0.70-1.93 for moderate was 1.72 95%CI 1.11-2.67, and severe was 2.87 95% CI 1.70-4.85, indicating that those with moderate to severe sleep apnea are at the highest risk of sudden death. (**Figure 5a-c**)

**Figure 5:**
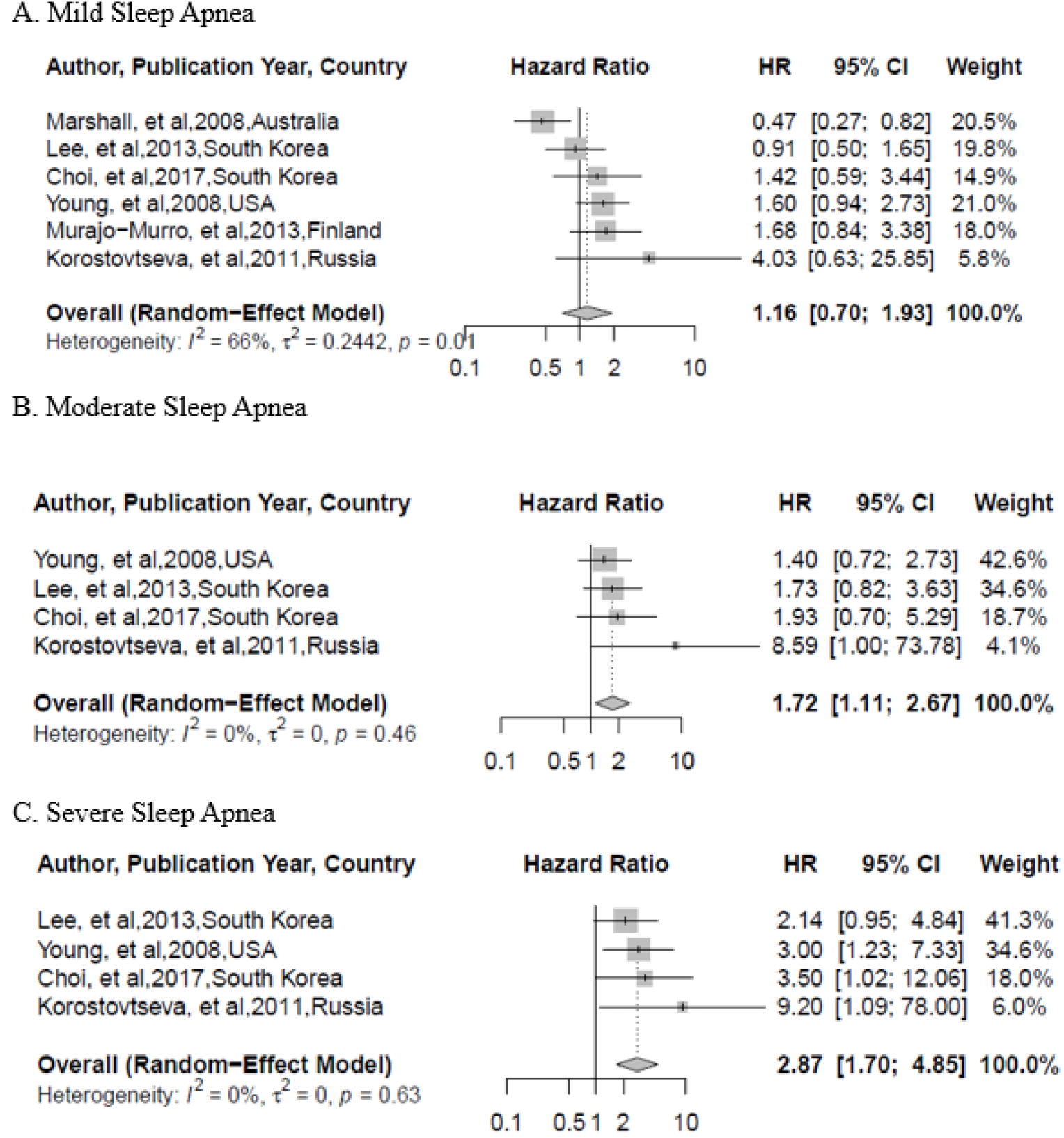
Dose response effects of sleep apnea on all-cause mortality. A. Mild sleep apnea was not associated with a significantly increased risk of mortality. B. Moderate sleep apnea increased the risk of mortality by 1.7 times (HR= 1.72, 95%CI 1.11-2.67). C. Severe Sleep apnea increased the risk of mortality by 2.9 times (HR=2.87, 95%CI 1.70-4.85).

### Sub-group analysis based on quality scoring

Of the 22 studies used for quantitative analysis, one had a quality score of 6, one had a quality score of 7, three had a score of 8 and seventeen had a score of nine (**Figure 6**). We performed sub-group analysis on studies with a score of nine compared to those with less than nine. In the studies with a score of 9, the relationship between OSA and mortality was significant (HR 1.91, 95%CI 1.50-244) but not in the studies with a lower quality score (HR 1.52, 95%CI 0.97-2.37). However the studies with higher scores were not statistically different from those with lower scores (p=0.37).

**Figure 6.**
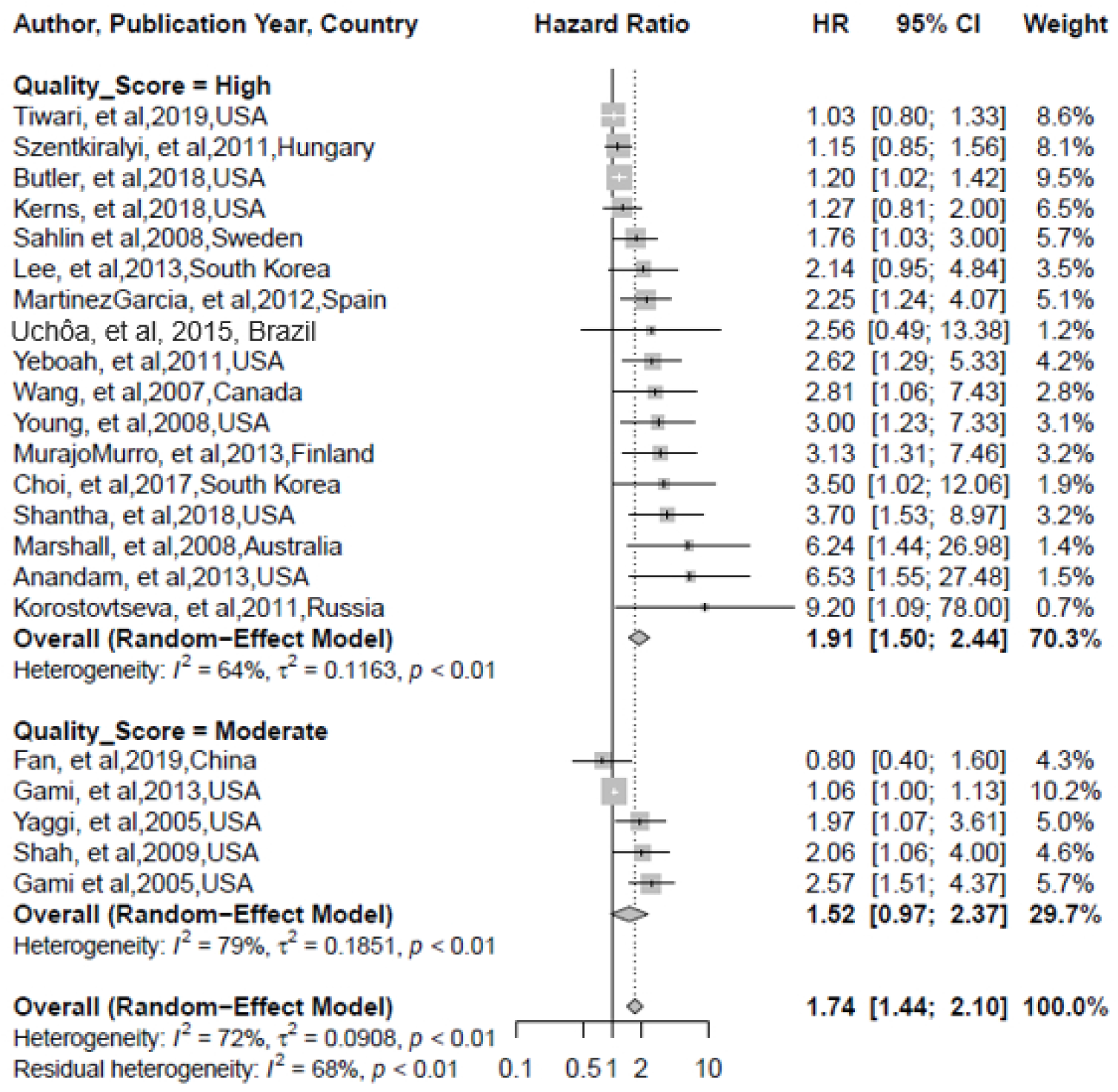
Subgroup analysis based on quality score of each paper. Although not statistically significant, publications with a higher quality scores, elicited a higher risk of mortality compared to those with a lower score.

## Discussion

To our knowledge, this is the first systematic review and meta-analysis to investigate all-cause mortality in patients with OSA. We screened six well-established databases (Pubmed-MEDLINE, OVID- HEALTH STAR, OVID-MEDLINE, Cochrane Library, Scopus, and Joana-Briggs Institute EBF Database) to identify all studies reporting the number of deaths in patients with obstructive sleep apnea. We screened 4,613 articles for inclusion criteria. After careful review, we included 28 studies. With this extensive search strategy, we discovered that (1) those with OSA displayed a hazard of sudden death that is 1.74 times higher than those without OSA, and (2) those with OSA displayed a hazard of cardiovascular-related mortality that is 1.94 times than those without OSA.

There is a large amount of evidence in research that supports the notion that OSA is associated with numerous cardiovascular conditions, including hypertension, coronary artery disease (CAD), congestive heart failure (CHF), arrhythmias, and more [41]. This association may be explained by the influence that the nervous system has on the sleep cycle in humans. OSA results in intermittent hypoxia and oxygen desaturation during sleep, which may cause over-arousal of the central nervous system (CNS) in order to increase airflow. The complex relationship between the sympathetic nervous system (SNS) and autonomic nervous system (ANS) causes a brief increase in both systolic and diastolic blood pressure (BP) during apneas [42-43]. The repeated exposure to an elevated BP continuously stresses the body. If this exposure is combined with a chronic cardiac condition, the wear on the body’s regulatory systems is even greater. Therefore, we hypothesize that constant stress on the body as a result of OSA may work to explain why the mortality rates are increased in this group.

In addition to this, patients with OSA also experience an increased level of stress on the biochemical systems in the body. Prior studies have explored the association between C-reactive protein (CRP) and obstructive sleep apnea [44]. C-reactive protein is known as an inflammatory marker. Research has found that patients with OSA have an increase in this protein, which simultaneously increases the oxidative stress they experience [44]. This chronic inflammation, which is widely prevalent in patients with OSA, is associated with endothelial cell injury. This is another hypothesis that may work to introduce some biological reasons for why mortality is increased in populations with OSA.

The methodology we employed allowed us to assess a large number of studies for inclusion. As a result of this, studies included in this meta-analysis were representative of many continents, including North America, Australia, Europe, Asia, and South America. With this being said, we were unable to identify any studies from Africa. There is a need for further research in these regions in order to identify if this association between OSA and mortality remains true. In addition to this, we only explored the association between cardiovascular mortality and sudden death in relation to OSA. Although the studies included in this review were comprehensive and thorough, they did not cover many other conditions. Factors such as obesity, diabetes, and others may be of interest to consider. Therefore, these conditions may not have been represented in this meta-analysis. Additional research should be conducted in order to explore if any other conditions can significantly increase the risk of death in patients with OSA.

## Conclusion

Individuals with OSA are at an increased risk for both cardiovascular mortality and sudden death. Treatments and interventions related to decreasing this risk and other adverse outcomes are necessary in order to optimize survival and quality of life (QOL).

## Data Availability

All data is available within table 1 and the figures

## Contributorship

AS, PS, ES, and VC conceived the study. AS, EH, and PS conducted the literature search. AS and PS completed data analysis. AS, EH, PS, and VC interpreted the data. AS, ES, and PS wrote the manuscript. All Authors agreed to the manuscript in its final form

## Patient and Public Involvement

Patients or the public were not involved in the preparation or dissemination of this manuscript

## Ethics Statement

This is a systematic review and meta-analysis and individual patient was not used. Therefore we did not need IRB or an ethics board approval.

## Data Sharing Statement

All data relevant to the study are included in the article or uploaded as supplementary information

---

**Table 1:**
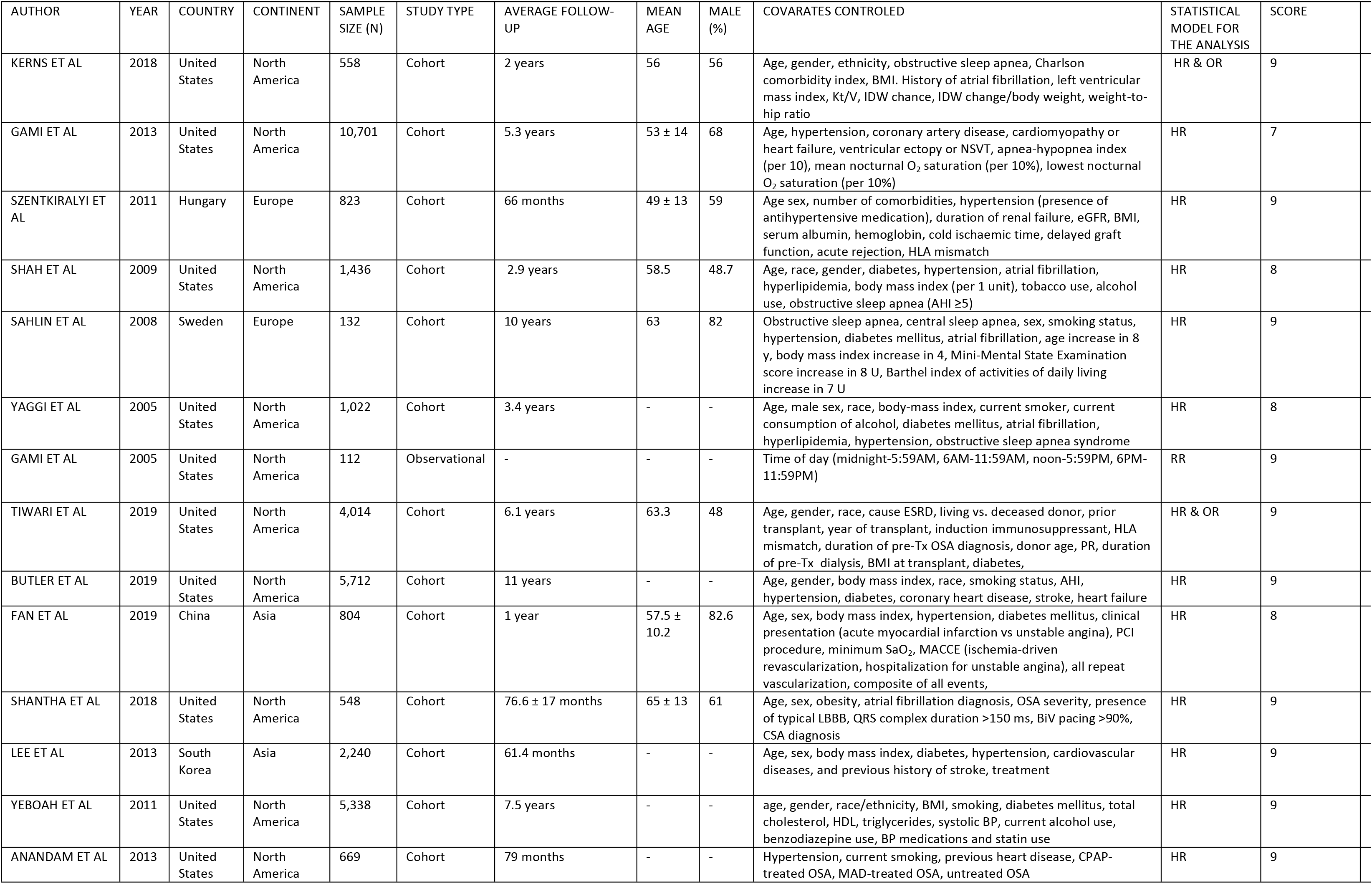
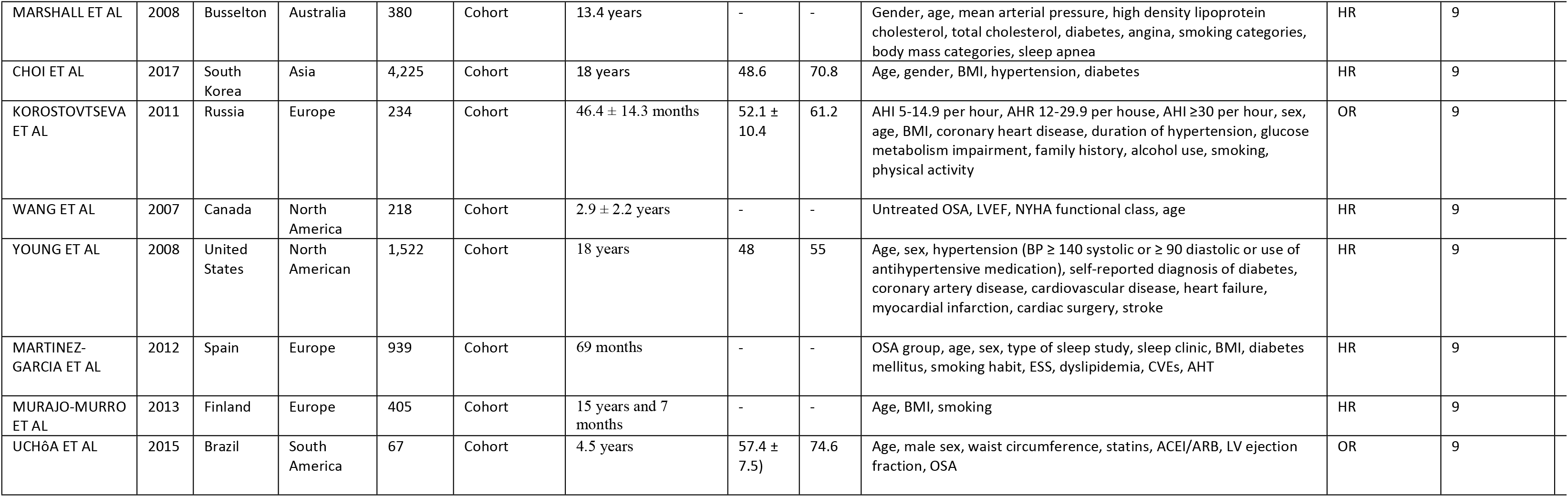
Studies of inclusion

